# Are Brain Responses to Emotion a Reliable Endophenotype of Schizophrenia? An Image-based fMRI Meta-analysis

**DOI:** 10.1101/2022.05.31.22275506

**Authors:** Anna M. Fiorito, André Aleman, Giuseppe Blasi, Josiane Bourque, Hengyi Cao, Raymond C. K. Chan, Asadur Chowdury, Patricia Conrod, Vaibhav A. Diwadkar, Vina M. Goghari, Salvador Guinjoan, Raquel E. Gur, Ruben C. Gur, Jun Soo Kwon, Johannes Lieslehto, Paulina B. Lukow, Andreas Meyer-Lindenberg, Gemma Modinos, Tiziana Quarto, Michael J. Spilka, Venkataram Shivakumar, Ganesan Venkatasubramanian, Mirta Villarreal, Yi Wang, Daniel H. Wolf, Je-Yeon Yun, Eric Fakra, Guillaume Sescousse

## Abstract

**Background:** Impaired emotion processing constitutes a key dimension of schizophrenia and a possible endophenotype of this illness. Empirical studies consistently report poorer emotion recognition performance in patients with schizophrenia as well as in individuals at enhanced risk of schizophrenia (“at risk”). fMRI studies also report consistent patterns of abnormal brain activation in response to emotional stimuli in patients, in particular decreased amygdala activation. In contrast, brain-level abnormalities in at-risk individuals are more elusive. We address this gap using an image-based meta-analysis of the fMRI literature.

**Methods:** fMRI studies investigating brain responses to negative emotional stimuli and reporting a comparison between at-risk individuals and healthy controls were identified. Frequentist and Bayesian voxel-wise meta-analyses were performed separately, by implementing a random effect model with unthresholded group-level T-maps from individual studies as input.

**Results:** Seventeen studies with a cumulative total of 677 at-risk individuals and 805 healthy controls were included. Frequentist analyses did not reveal significant differences between at-risk individuals and healthy controls. Similar results were observed with Bayesian analyses, which provided strong evidence for the absence of meaningful brain activation differences across the entire brain. Region of interest analyses specifically focusing on the amygdala confirmed the lack of group differences in this region.

**Conclusions:** These results suggest that brain activation patterns in response to emotional stimuli are unlikely to constitute a reliable endophenotype of schizophrenia. We suggest that future studies rather focus on impaired functional connectivity as an alternative and promising endophenotype.

## Introduction

Schizophrenia is a chronic and profoundly disabling psychiatric disorder that has a significant impact on patients’ well-being(1). A core feature of the disorder is impairment in emotion processing, in particular poorer emotion recognition performance. Indeed, numerous studies show that patients with schizophrenia have deficits in emotion perception, particularly during facial emotion processing(2,3). Moreover, these deficits may play a key role in schizophrenia’s etiopathogenesis(4). It has been proposed that vulnerability to schizophrenia itself could be linked to an overall tendency to experience more negative affect(5). Behavioral deficits have also been accompanied by functional brain abnormalities: several meta-analyses have described atypical blood oxygenation level-dependent (BOLD) activation in patients with schizophrenia compared with healthy controls during emotion perception(6–12). The most consistent finding is a hypoactivation of the amygdala during emotion processing(6–11).

Individuals at enhanced risk of schizophrenia (hereafter called “at risk”) tend to show similar but more subtle behavioral deficits when processing emotions(13). Meta-analyses show moderate abnormalities in emotion recognition in healthy first-degree relatives(14,15) (who share part of the genetic vulnerability and therefore are considered at familial risk(16)) and individuals at clinical high risk(17) (CHR, who are considered at risk based on clinical criteria(18)). Evidence for these abnormalities is particularly notable for negative emotions(15). In a similar vein, individuals with high schizotypy traits based on self-report questionnaires (psychometric risk(19)) experience more difficulties in recognizing emotions than individuals with low schizotypy traits(20). Therefore, it has reasonably been proposed that these deficits could represent a candidate endophenotype of schizophrenia(21), a measurable component along the pathway between the disease and the distal genotype, but less readily observed in at-risk individuals(22).

Neuroimaging measures are considered as particularly informative endophenotypes(23,24) because they may be more stable than behavioral performance which varies considerably throughout the course of the illness(21). However, there is a relative paucity of results from neuroimaging studies investigating emotion processing in at-risk individuals, and observed differences in activation-based measures are inconsistent. Moreover, individual studies have been characterized by relatively modest sample sizes, hence providing limited statistical power. Several studies have relied on region of interest approaches, thus failing to provide information on whole-brain differences in brain activation(25,26). Some studies have reported decreased activation in the amygdala(27–29), partially consistent with the literature on schizophrenia. However, there are several concurrent reports of hyperactivation of the amygdala(30,31), or of null results(32–34). Meta-analyses are ideally suited for disambiguating such confusion, and two of them have actually been performed in an attempt to quantify brain abnormalities during the processing of emotional stimuli in healthy first-degree relatives(35) and CHR individuals(36). While no significant differences were found in the latter group(36), widespread clusters of hyperactivation were found in healthy first-degree relatives(35). However, due to the small number of included studies (N=4(35,36)), these meta-analyses had limited statistical power, motivating the need of a more powerful meta-analysis.

Indeed, this uncertainty clouds our ability to properly assess the value of activation-based differences during emotion processing as reliable endophenotypes in the schizophrenia diathesis(37). Accordingly, our main objective was to clarify whether at-risk individuals exhibit abnormal brain responses to negative emotional stimuli compared with healthy controls. To investigate this question, we performed an image-based meta-analysis, using functional neuroimaging data acquired across multiple studies. Whole-brain frequentist analyses, implemented using a random-effect model in the Seed-based d Mapping (SDM) software, and Bayesian analyses, were performed. This latter approach allowed us to quantify evidence in favor of the null hypothesis (absence of a group difference) and the alternative hypothesis (existence of a group difference). In both statistical frameworks, we first examined brain responses to negative emotional stimuli versus non-emotional stimuli (i.e., neutral stimuli, control condition, or implicit baseline) separately in at-risk individuals (i.e., familial risk, CHR, psychometric risk) and healthy controls (within-group meta-analysis), and then in at-risk individuals versus healthy controls (between-group meta-analysis). Finally, in light of previous findings in patients, activation differences in the amygdala were specifically investigated via region of interest (ROI) analyses.

## Methods and Materials

### Inclusion of studies

The meta-analysis was conducted in accordance with the Preferred Reporting Items for Systematic Reviews and Meta-analyses guidelines (PRISMA, 2020, see Table S1 in the Supplement for the checklist). The protocol for this meta-analysis was not pre-registered. We performed a comprehensive literature search using PubMed and Web of Science. The search (which identified records published until 15^th^ April 2020) used a combination of terms constructed according to 4 stems relating to (a) schizophrenia (schizophren*, psychosis), (b) at-risk individuals (relatives, first-degree, siblings, twins, brothers, sisters, offspring, parents, high risk, ultrahigh risk, ultra-high risk, genetic risk, clinical risk, prodromal, ARMS, at-risk mental state) (c) neuroimaging (neuroimaging, functional Magnetic Resonance Imaging, fMRI), and (d) emotions (emotion*, affect, mood). Moreover, an inspection of the reference list of included studies was performed in order to find articles that were not identified through the database search.

Inclusion criteria were the following: studies that (a) were written in English; (b) reported a comparison between at-risk individuals (i.e., familial risk, CHR, psychometric risk) and healthy controls; (c) used task-based functional magnetic resonance imaging (fMRI) with an emotion perception element, and (d) included negatively valenced emotional stimuli. We focused on negative stimuli because this class of stimuli is more heavily represented in emotional tasks and appears to induce stronger behavioral impairments in individuals at enhanced risk compared with positive stimuli(15).

The following exclusion criteria were applied: studies that (a) did not report an emotional versus non-emotional contrast, and for which the authors could not provide the contrast when queried; (b) employed a paradigm non-specific to emotion processing (e.g., conditioning paradigm, Theory of Mind paradigm); (c) did not mainly use visual emotional stimuli of faces or scenes in their protocol; (d) primarily employed positively valenced emotional stimuli; (e) did not use unique populations (i.e., used data from another included study), since inclusion of results that are not statistically independent can inflate Type I error(38); (f) could not provide a whole-brain analysis; (g) could only provide a general F-statistic instead of direct comparisons between groups; (h) were published conference papers, and (i) were systematic reviews or meta-analyses.

The literature search yielded a total of 308 unique papers (see Figure 1). Initial screening based on title or abstract was performed (by AF). After these exclusions, 45 full articles were independently screened (by AF and GS) to be assessed for eligibility. Disagreements were resolved through discussion and mediation by a third author (EF). Our strict application of inclusion and exclusion criteria filtered the analysis to a total of 26 unique papers. The corresponding authors on these 26 papers were contacted to obtain unthresholded group-level T-maps, associated with within- and between-groups effects. Following this correspondence, we included data for 16 studies. Data from the remaining studies could not be included(30,39–47) either because corresponding authors were not responsive (after multiple reminders), the required data were lost and/or impossible to retrieve, or the data was found to be dubious (see Supplementary Methods for more details on study exclusion). Our correspondence also revealed the existence of a yet unpublished study meeting all inclusion criteria (Shivakumar, in preparation(48)), bringing the number of included studies to a total of 17.

**Figure 1.**
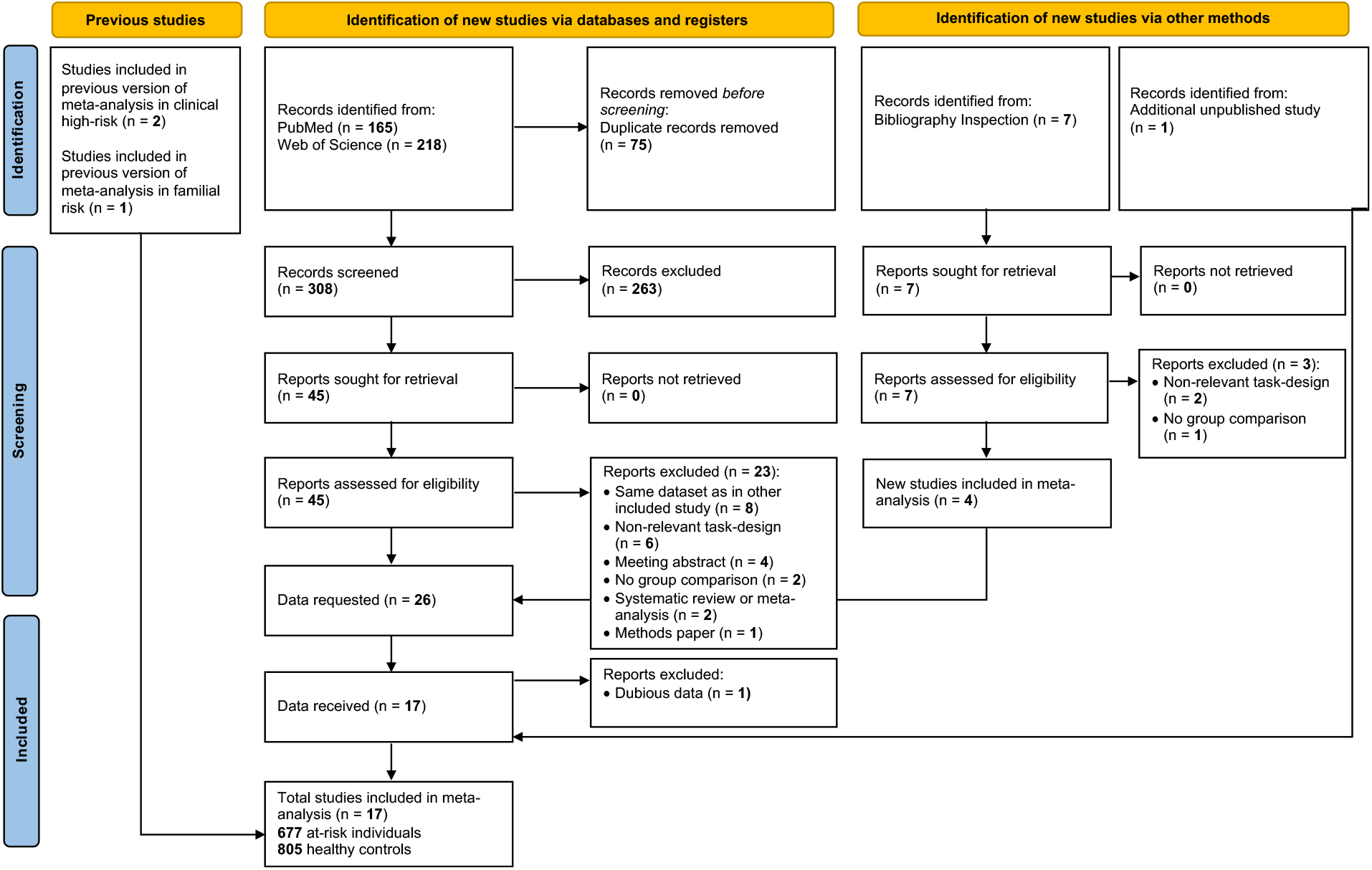
Flow Chart outlining the selection procedure of studies

### Study characteristics

The included studies encompassed two types of tasks: a) “implicit” tasks, which employed emotional stimuli while using instructions with a non-emotional focus (e.g., passive viewing of emotional images, age or gender recognition), and b) “explicit” tasks, in which instructions specifically require to focus on the emotional component of stimuli (e.g., emotion recognition, emotion regulation, emotion evaluation, rating of emotional arousal, matching of emotional faces). Details regarding the task design and included contrasts for each study are reported in Table 1. Further details (mean age, percentage of male participants, field strength) are reported in Table S2 in the Supplement. Following recommendations from the latest version of SDM, we only included a single contrast per study (see Supplementary Methods for selection criteria).

**Table 1.** Included studies

| Reference | Definition of at risk | Number of Participants | Task Used | Explicit/ Implicit task | Type of stimuli | Contrast included |
| --- | --- | --- | --- | --- | --- | --- |
| Bourque et al., 2017(49) | Psychometric | At risk (27); HC (135) | Passive viewing | Implicit | Faces | Angry Stimuli > Control Condition |
| Cao et al., 2016(50) | Familial (mixed first-degree) | At risk (58); HC (94) | Emotion matching | Explicit | Faces | Negative Stimuli > Control Condition |
| de Achával et al., 2012(51) | Familial (siblings) | At risk (14); HC (14) | Gender recognition | Implicit | Faces | Positive and Negative Stimuli > Implicit baseline<sup>a,b</sup> |
| Diwadkar et al., 2012(25) | Familial (offspring) | At risk (17); HC (25)<sup>c</sup> | Emotion recognition | Explicit | Faces | Negative Stimuli > Neutral Stimuli<sup>a</sup> |
| Modinos et al., 2010(52) | Psychometric | At risk (17); HC (17) | Emotion regulation | Explicit | IAPS (complex scenes of people) | Negative Stimuli > Neutral Stimuli<sup>a</sup> |
| Modinos et al., 2015(28) | CHR | At risk (18); HC (20) | Emotional arousal rating | Explicit | IAPS (complex scenes of people, animals, and objects) | Negative Stimuli > Neutral Stimuli<sup>a</sup> |
| Modinos et al., 2017(34) | Psychometric | At risk (21); HC (22) | Emotional arousal rating | Explicit | IAPS (complex scenes of people, animals, and objects) | Negative Stimuli > Neutral Stimuli<sup>a</sup> |
| Park et al., 2016(27) | Familial (mixed first-degree) | At risk (20); HC (17) | Gender recognition | Implicit | Faces | Fearful Stimuli > Implicit baseline |
| Pulkkinen et al., 2015(53) | Familial (offspring) | At risk (51); HC (52) | Passive viewing | Implicit | Faces | Fearful Stimuli > Control Condition |
| Quarto et al., 2018(54) | Familial (siblings) | At risk (36); HC (56) | Gender recognition | Implicit | Faces | Fearful Stimuli > Neutral Stimuli<sup>a</sup> |
| Rasetti et al., 2009(55) | Familial (siblings) | At risk (29); HC (20) | Emotion matching | Explicit | Faces | Negative Stimuli > Control Condition |
| Spilka et al., 2015(32) | Familial (mixed first-degree) | At risk (27); HC (27) | Passive viewing | Implicit | Faces | Negative Stimuli > Neutral Stimuli<sup>a</sup> |
| van der Velde et al., 2015(33) | CHR | At risk (15); HC (16) | Emotion regulation | Explicit | IAPS (complex scenes of people) | Negative Stimuli > Neutral Stimuli |
| Shivakumar et al., in preparation (48) | Familial (mixed first-degree) | At risk (13); HC (15) | Emotion matching | Explicit | Faces | Negative Stimuli > Control Condition |

| Reference | Definition of at risk | Number of Participants | Task Used | Explicit/Implicit task | Type of stimuli | Contrast included |
| --- | --- | --- | --- | --- | --- | --- |
| Wang et al., 2018(29) | Psychometric | At risk (34); HC (30) | Emotion recognition | Explicit | Faces | Fearful Stimuli > Neutral Stimuli |
| Wolf et al., 2011(56) | Familial (mixed first-degree) | At risk (20); HC (25) | Emotion recognition | Explicit | Faces | Negative Stimuli > Neutral Stimuli<sup>a</sup> |
| Wolf et al., 2015(31) | CHR | At risk (260); HC (220) | Emotion recognition | Explicit | Faces | Fearful Stimuli > Neutral Stimuli<sup>a</sup> |

Abbreviations: HC, Healthy Controls; CHR, clinical high risk; IAPS, International Affective Picture System
<sup>a</sup> The included contrast was not the one originally reported in the article but a new generated one.
<sup>b</sup> Note that we included one study with a contrast involving positively valenced stimuli. Nevertheless, around 70% of the stimuli represented negative emotions. We therefore considered this study as largely representing negative emotions and we refer to the general contrast included in this meta-analysis as a negative emotional stimuli versus non emotional stimuli contrast.
<sup>c</sup> This number corresponds to the dataset sent by the authors which is slightly different from the dataset reported in the paper. This is because the authors retrieved the latest available dataset

#### Analyses

We ran an image-based meta-analysis using statistically unthresholded T-maps provided by authors. This approach is more sensitive compared with more common coordinate-based meta-analyses(57,58), as it accounts for effect sizes and is also sensitive to moderate subthreshold effects that are consistent across studies(59).

For the sake of homogeneity, some of the included contrasts were not those reported in the original articles, but new contrasts generated by the authors at our request based on their methodological design (see Table 1).

### Whole-Brain Frequentist Analyses

Frequentist meta-analyses were performed using the latest version of SDM based on Permutation of Subject Images (SDM-PSI, version 6.21(60)). Traditional meta-analysis methods (e.g., Activation Likelihood Estimation, ALE, and Multilevel Kernel Density Analysis, MKDA) test for spatial convergence, evaluating in each voxel whether studies found activation more often in one group compared with another. In contrast, SDM-PSI employs a new algorithm that tests whether activation in each voxel is significantly different between groups(60). This approach has been argued to be more robust and has several advantages: the interpretation of results is more straightforward and is not based on spatial assumptions that could be violated, and voxel-level significance is independent of the distribution of activation across the brain. Moreover, this method accounts for both activations and deactivations, allowing opposite findings to potentially cancel each other. Finally, the method employs a threshold-free cluster enhancement (TFCE) approach to address the multiple comparison issue, which compared with other thresholding techniques has the benefit of not defining an arbitrary cluster-forming threshold(61).

We included contrasts broadly depicting a comparison between the perception of negative emotional stimuli versus non-emotional stimuli (i.e., neutral stimuli, control condition, or implicit baseline). We initially examined these contrasts separately in at-risk individuals and healthy controls (within-group meta-analysis). Of the 17 studies included in the meta-analysis, one could not provide within-group T-maps and therefore was excluded from this analysis(50). Thus, the within-group meta-analysis included 16 studies, cumulating 619 at-risk individuals and 711 healthy controls. Then, the same contrasts were examined in at-risk individuals versus healthy controls (between-group meta-analysis). 17 studies were included in this analysis for a total of 677 at-risk individuals versus 805 healthy controls.

Results were corrected for multiple comparisons across the whole brain using a TFCE-corrected threshold of p_TFCE_<0.05.

### Whole-Brain Bayesian Analyses

Bayesian analyses were performed with the BayesFactorFMRI(62,63) toolbox to quantify evidence in favor of the null and alternative hypotheses. This toolbox employs R and Python code to perform Bayesian voxel-wise meta-analysis of T-maps. The output is a meta-analytical map storing a Bayes factor (BF) for each voxel. The Bayes factor is the ratio of the likelihood of the data under the alternative hypothesis (H_1_, hypothesizing a meaningful effect in a within- or between-group analysis) and the likelihood of the same data under the null hypothesis (H_0_, hypothesizing the absence of a meaningful effect in a within- or between-group analysis). BF_10_ quantifies the evidence in favor of H_1_ compared with H_0_, while BF_01_ (=1/BF_10_) quantifies the evidence in favor of H_0_ compared with H_1_. Conventionally a BF_10_ (or BF_01_) that exceeds the threshold of 3 represents moderate evidence in favor of H_1_ (or H_0_), while a Bayes factor between 10 and 30 represents strong evidence(64). Results are thus reported at a threshold of BF>3.

The BayesFactorFMRI toolbox only computes Bayes factors in voxels that are covered by all included studies (similar to second-level analyses performed in SPM or FSL). Therefore, one study with partial brain coverage(56) (see Supplementary Methods) had to be excluded from these analyses in order to retain information in a substantial part of the brain covered by all the remaining studies. The approach was similar to the initial frequentist analyses: we first performed within-group analyses (n=15 studies), examining the negative emotional versus non-emotional contrast in at-risk individuals (n=599) and healthy controls (n=686) before performing the between-group analyses (n=16 studies) in at-risk individuals (n=657) versus healthy controls (n=780).

### ROI Analyses (frequentist and Bayesian)

Given the key role of the amygdala in emotion processing, and previous findings in patients with schizophrenia(6–11), the bilateral amygdala was selected for region of interest based analyses. The amygdala was anatomically defined using the Melbourne Subcortex Atlas(65).

Averaged effect sizes and associated variances were extracted separately for the left and right amygdala for each study using SDM-PSI. These values were used to perform frequentist and Bayesian meta-analyses as well as to generate forest plots and contour-enhanced funnel plots.

Frequentist ROI meta-analyses were performed with SDM-PSI. Bayesian ROI meta-analyses were performed with R (version 4.1.2) by fitting a random-effect Bayesian model (using the *brms* package(66)), and defining weakly informative priors for the mean (normal distribution centered at 0 with a variance of 1) and variance (Half-Cauchy distribution with peak at 0 and scaling parameter of 0.5) of the pooled true effect as suggested by Harrer et al., 2021(67).

## Results

Unthresholded whole-brain maps of within- and between-group meta-analyses are available on NeuroVault(59), for both frequentist and Bayesian analyses (https://neurovault.org/collections/CRLVVOUU/).

### Whole-Brain Frequentist Analyses

Across included studies (n=16), the meta-analyses of within-group T-maps in healthy controls and in at-risk individuals revealed significant activations in several brain regions typically involved in emotion processing, such as bilateral amygdala, thalamus, and inferior frontal gyrus (Figure 2, see Table S3 in the Supplement for MNI coordinates). The pattern of activation and deactivation was remarkably similar between healthy controls and at-risk individuals.

**Figure 2.**
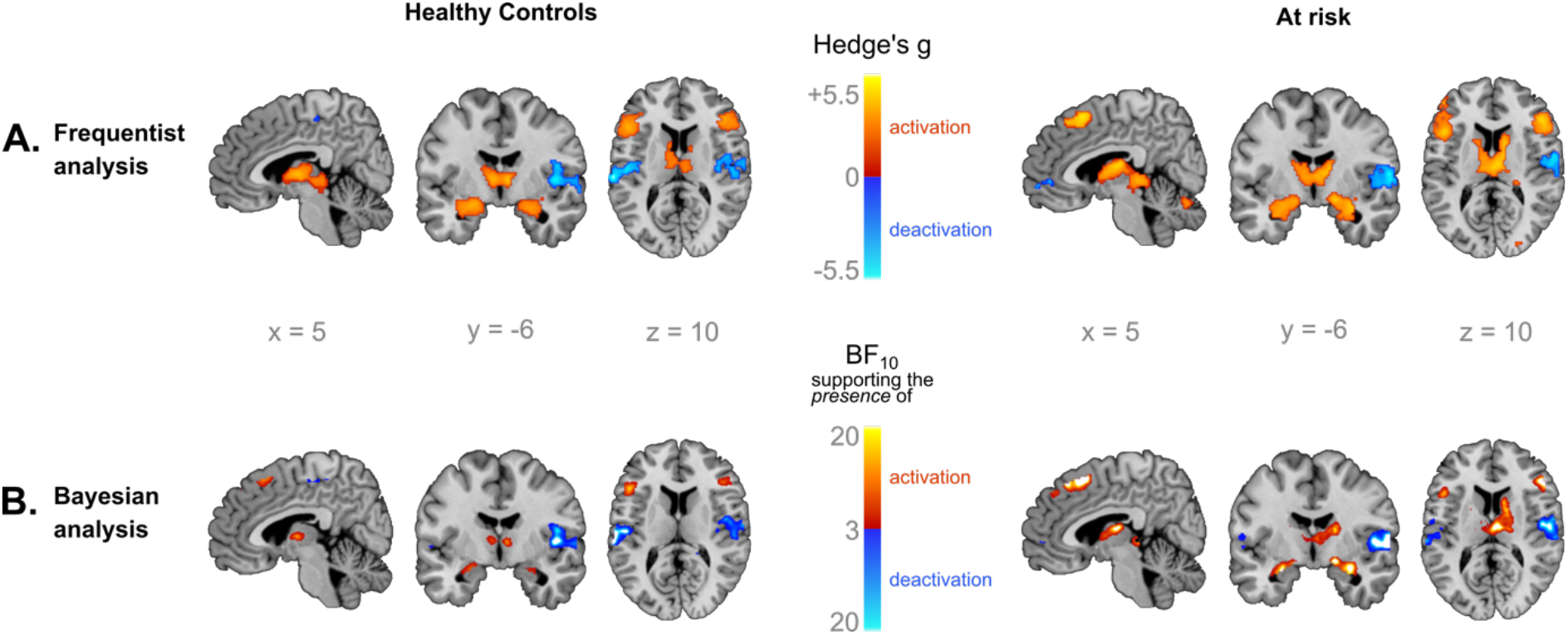
Within-group meta-analyses of negative emotion processing. Brain regions showing activation (in red) and deactivation (in blue) during negative emotion processing as revealed by frequentist (**A**.) and Bayesian (**B**.) analyses, for healthy controls and at-risk individuals. Regions typically involved in emotion processing were activated (e.g., bilateral amygdala, thalamus, inferior frontal gyrus). Note that results were consistent across statistical approaches, and that the patterns of activation and deactivation were remarkably similar in healthy controls and at-risk individuals. Functional maps are overlaid on the Colin 27 anatomical template. Frequentist analyses are thresholded at p_TFCE_<0.05 and Bayesian analyses are thresholded at BF_10_>3.

The whole-brain between-group meta-analysis (n=17 studies) revealed no significant group differences at p_TFCE_<0.05.

### Whole-Brain Bayesian Analyses

Results of within-group Bayesian meta-analyses (n=15 studies) largely reproduced the activation pattern revealed by frequentist analyses, showing activation in bilateral amygdala, thalamus, and inferior frontal gyrus (Figure 2).

The between-group Bayesian meta-analysis (n=16 studies) revealed two small clusters showing moderate evidence in favor of increased activation in at-risk individuals compared with healthy controls. These clusters were observed in the left superior frontal gyrus (x=-6, y=34, z=46, BF_10_=4.78, voxel size=10) and right peri-hippocampal white matter (x=20, y=-12, z=-10, BF_10_=7, voxel size=6) (BF_10_, see Figure 3 in red). No region showed evidence in favor of a decreased activation in at-risk individuals compared with healthy controls (BF_10_, see Figure 3 in blue). Instead, evidence in favor of the absence of differences in brain activation in at-risk individuals compared with healthy controls was found across almost the entire brain, with BF_01_ reaching values up to 30, indicating strong evidence in favor of H_0_ (BF_01_, see Figure 3 in green).

**Figure 3.**
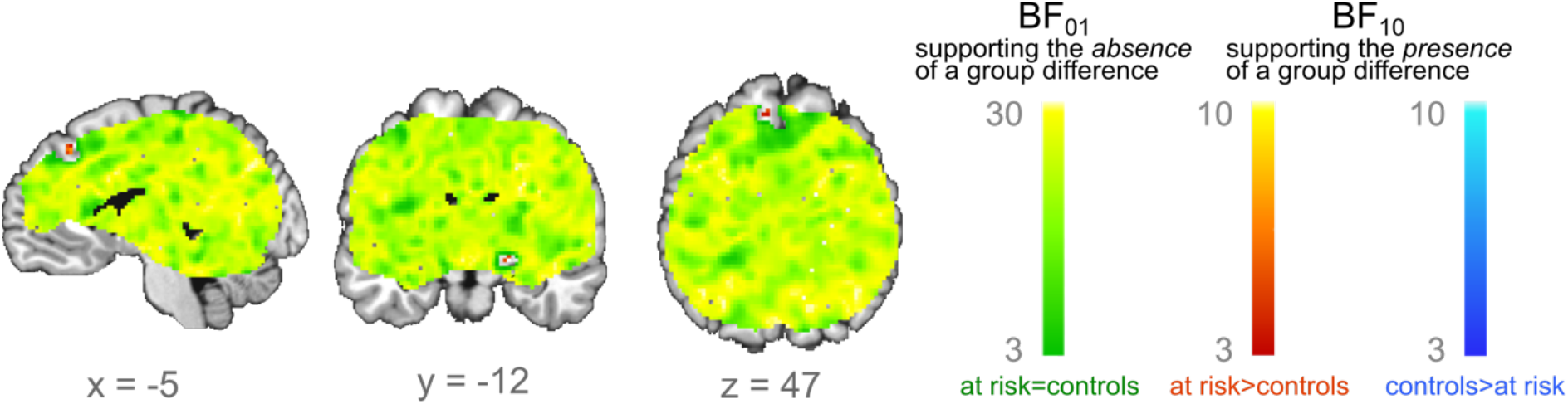
Between-group Bayesian meta-analysis of negative emotion processing. Voxels with moderate evidence supporting hyperactivation in at-risk individuals compared with healthy controls are depicted in red (BF_10_>3). Voxels with at least moderate evidence supporting a hypoactivation in at-risk individuals compared with healthy controls are depicted in blue (BF_10_>3). Voxels with at least moderate (or higher) evidence supporting the absence of differences in at-risk individuals compared with healthy controls are depicted in green (BF_01_>3). Only two small clusters showed moderate evidence (i.e., 3<BF_10_<10) in favor of hyperactivation in at-risk individuals compared with healthy controls. In contrast, strong evidence (i.e., most BF_01_>10) in favor of the absence of group differences was found almost across the whole brain. Functional maps are overlaid on the Colin 27 anatomical template. Results are thresholded at BF>3.

### ROI Analyses (Frequentist and Bayesian)

Frequentist meta-analysis in amygdala ROIs revealed no significant group differences in either the left (g=0.00, p=0.52) or right (g=0.02, p=0.58) hemisphere (Figure 4A).

**Figure 4.**
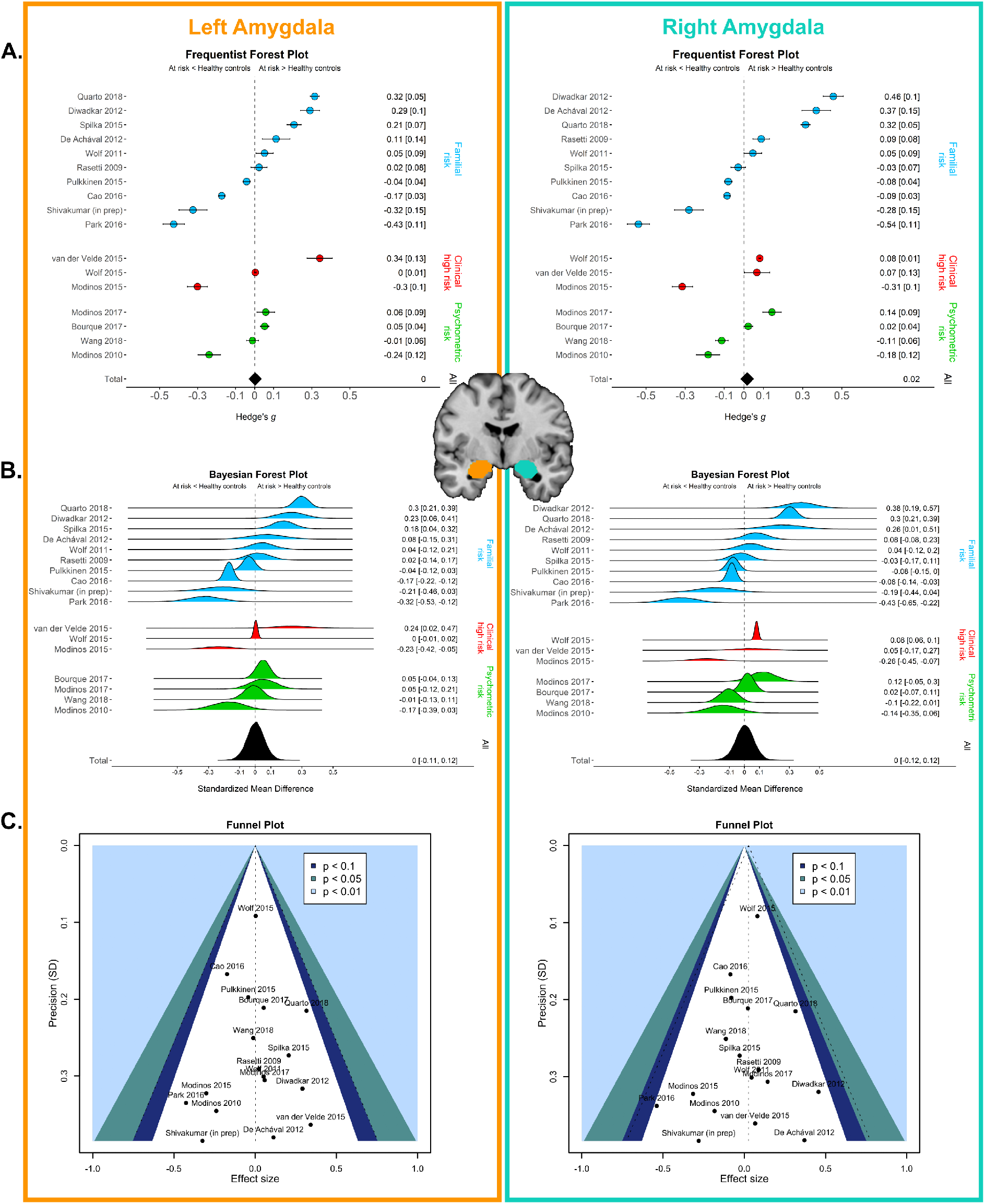
ROI meta-analyses of negative emotion processing. Graphical representations of values extracted from left and right amygdala: **A**. Frequentist forest plot depicting the mean ± variance of effect sizes for group comparison. The black diamond represents the overall effect size; **B**. Bayesian forest plot depicting Bayesian estimates of the true effect size and posterior distribution of each study for group comparison. Posterior distribution depicted in black represents the Bayesian meta-estimate and posterior distribution of the overall effect size. Note that for both Bayesian and frequentist forest plots, distinguishing subgroups as a function of the definition of “at risk” did not seem to impact the results; **C**. Contour-enhanced funnel plot: the dotted part depicts the overall effect size and 95% confidence interval around this overall effect size, while the colored areas represent different levels of statistical significance (p<0.1, p<0.05, p<0.01, corresponding to 90%, 95%, and 99% confidence interval around an effect size of 0)(68). Symmetrical funnel plots for both the left and right amygdala suggest no evidence for publication bias, which is further strengthened by all studies lying in the area of statistical non-significance (p>0.1).

Bayesian meta-analysis found strong evidence for the absence of group differences between at-risk individuals and healthy controls in left (Standardized Mean Difference=0, [95%CI -0.11–0.12]; BF_01_=18.2) and right amygdala (Standardized Mean Difference=0, [95%CI -0.12–0.12]; BF_01_=17.31) (Figure 4B).

### Heterogeneity, Publication Bias and Robustness

In additional sensitivity analyses (see Supplementary Methods for details) we show that results did not differ according to the definition of 1) “at risk of schizophrenia” (i.e., familial risk, CHR, or psychometric risk), 2) type of contrast (i.e., negative emotional stimuli versus neutral stimuli, or versus control condition/implicit baseline), 3) type of task (implicit or explicit), or 4) type of emotional stimuli (i.e., when restricting analyses to those studies only including faces). We also show that our data have low heterogeneity (see Figure S1 in the Supplement) and showed no obvious sign of publication bias (Figure 4C), and that the results were not driven by outlier studies.

## Discussion

Our meta-analysis aimed to investigate whether brain activation in response to negative emotional stimuli could be considered as an endophenotype of schizophrenia. Results show that healthy controls and individuals at risk of schizophrenia present a remarkably similar pattern of brain activation in response to processing negative emotional stimuli. These similarities were observed in salient regions of the emotion processing network including the amygdala, thalamus, and inferior frontal gyrus. In line with this observation, the between-group meta-analysis did not reveal group differences. Notably, not only did the frequentist meta-analysis failed to reveal significant differences between healthy controls and at-risk individuals, but the Bayesian meta-analysis provided strong support (i.e., BF_01_>10) for the absence of such group differences across the entire brain. Two small clusters showed hyperactivation in at-risk individuals, but these were located in regions irrelevant to the process at stake, were only observed in Bayesian analyses, and were associated with only moderate evidence (i.e., BF_10_<10). We thus prefer to refrain from speculating about these results, which do not appear to be robust. Finally, ROI analyses specifically focusing on the amygdala confirmed the absence of between-group activation differences. This null result is particularly notable, because the amygdala plays a primary role in emotion processing, and has shown blunted responses in patients with schizophrenia in multiple meta-analyses(6–11). Given the combination of frequentist and Bayesian approaches and the careful curation of the included datasets, we believe that our results are particularly strong and reliable. The inescapable inference is that in at-risk individuals, brain responses to negative emotional stimuli are too variable to be considered as a reliable endophenotype of schizophrenia. This observation is consistent with a previous meta-analysis of brain responses to emotional stimuli in CHR individuals(36), which did not find any differences with healthy controls.

Our strategy has the merit of bringing together several definitions that are commonly used to identify at-risk individuals (i.e., familial, CHR, psychometric), exploring the vulnerability spectrum of schizophrenia. Yet, these different definitions of risk (either based on genetic vulnerability, sub-threshold clinical symptoms, or high schizotypy traits) as well as differences in transition rates(69–71) may increase heterogeneity and consequently mask subtle but reliable group differences. In order to identify potential variability across definitions, we therefore conducted separate subgroup analyses (see Supplementary Methods). None of these analyses revealed significant results. Nevertheless, it is important to emphasize that some diagnostic uncertainty and heterogeneity still remain within each subgroup. This clinical variability introduces some additional variance that may diminish our ability to observe more subtle group differences. The use of mega-analytical methods employing subject-level data is a promising development to address this issue(58). Also, moving away from categorical to more dimensional definitions of mental health conditions should allow to better take into account this variability, by improving the granularity of nosological descriptions.

Moreover, while this meta-analysis focuses on emotion perception, emotion processing consists of multiple components such as experience, regulation, and expression. Examined individually, such components may be able to show robust association with schizophrenia vulnerability, allowing for refined candidate endophenotypes. For instance, there is some evidence of brain abnormalities in emotion regulation in patients with schizophrenia(44,72), which are also reported in at-risk individuals(33,52).

Finally, it should be noted that the amygdala, a key region in emotion processing, is particularly sensitive to susceptibility artifacts, leading to BOLD signal dropout. As a result, some of the included studies had only partial coverage of the amygdala. This observation calls for optimized acquisition protocols in future studies to circumvent this issue (see Supplementary Methods for a more detailed discussion).

Despite these limitations, we believe that the cutting-edge methodology of our meta-analysis provides strong empirical support for the results. First, we performed an image-based meta-analysis, which considers effect sizes from individual voxels across the entire brain as opposed to coordinate-based meta-analyses (such as ALE and MKDA) that rely only on peak localization(57,58). This difference bestows our approach with increased sensitivity. Second, we contacted authors of selected studies and solicited originally collected data with specific contrasts based on their methodological design. It is notable that, in addition to maximizing homogeneity across studies, this strategy is less susceptible to publication bias; indeed, both positive and negative results are collected, and studies reporting only ROI analyses are not systematically excluded, thus eliminating a potential bias(73). Third, we performed both within- and between-group analyses. In addition to confirming the activation of regions involved in emotion processing, the within-group meta-analyses revealed similar brain activation patterns in healthy controls and at-risk individuals. This similarity provides a straightforward and logical explanation for the absence of group differences in the between-group meta-analyses. Fourth, the Bayesian approach reproduced the results of the frequentist analyses, even when it was not subject to the typical limitations of the latter. Such convergence provides quantitative evidence in favor of the null hypothesis(74). Finally, this meta-analysis follows the recently updated PRISMA guidelines(75), ensuring the greatest level of transparency(76).

In conclusion, this meta-analysis reveals that individuals at risk of schizophrenia do not show clear abnormalities in brain activation during negative emotion perception. This finding has both clinical and theoretical implications for schizophrenia. In general, it is unclear whether emotional deficits are involved in schizophrenia etiopathogenesis, and therefore are present before the onset of the illness, or should be considered as a consequence of the disease itself (for instance, due to chronic effects of the disease, medication treatment, or physiological adaptation secondary to the illness). While it is well-established that brain responses to emotions are blunted in patients with schizophrenia, investigating these responses in at-risk individuals brings us a step closer to elucidating their role, helping to differentiate cause from consequence. In this meta-analysis, we reported an absence of group differences between at-risk individuals and healthy controls. This suggests that blunted brain responses to emotions are more likely to represent a consequence of the illness, i.e. a state marker which manifests itself only when the illness is active. Following this reasoning, abnormal brain responses to emotions are unlikely to be a useful endophenotype with the potential to help clinicians detect individuals at risk of schizophrenia.

It should also be noted that the endophenotype reasoning posits a continuum between the genetic risk and the expression of the abnormal trait. Yet, recent literature confirms that schizophrenia risk hinges on the interaction between genetic and environmental factors (77). Thus, the nonappearance of abnormal brain responses to emotions observed here in at-risk individuals could be explained by the absence, in the medical history of these subjects, of the environmental conditions necessary to reveal the phenotype variation.

Recent work has proposed that the blunted limbic responses observed in patients with schizophrenia during emotion processing could be at least partially explained by enhanced reactivity to neutral stimuli(12). Based on this observation, one might speculate that the neurotypical brain responses found in at-risk individuals might reflect a “downward normalization” of brain responses towards neutral stimuli, and not just an “upward normalization” of brain responses to emotional stimuli.

Crucially though, it should be emphasized that emotions are processed by a system of distributed and interconnected brain structures and that functional connectivity during emotion processing has been shown to be impaired in schizophrenia(78,79). Interestingly, a few studies investigating brain connectivity during emotion processing have also reported differences in individuals with familial risk(25,50,54), CHR(41) and psychometric risk(52) compared with healthy controls, mostly showing a blunted connectivity between limbic and frontal regions. In the framework of a chronic and poor-outcome disorder such as schizophrenia, endophenotypes are considered essential to help clinicians identify subjects at risk of schizophrenia and inform preventive approaches. Results of the present meta-analysis may prove useful to guide future research, and we suggest that future studies focus on the role of disrupted cortico-limbic coupling among at-risk individuals as a potential candidate endophenotype of schizophrenia.

## Data Availability

Unthresholded whole-brain maps of within- and between-group meta-analyses are available on NeuroVault at https://neurovault.org/collections/CRLVVOUU/

## Acknowledgment Section

AF and GS have full access to all the data in the study and take responsibility for the integrity of the data and the accuracy of the data analysis.

Concept and design: AF, GS, EF.

Acquisition (sharing data and/or performing additional analyses): All authors but AF, GS, EF.

Analyses and interpretation of data: AF, GS, EF.

Statistical analyses: AF, GS, EF.

Drafting of the manuscript: AF, GS, EF.

Critical revision of the manuscript for important intellectual content: All authors.

Obtained funding: EF.

Authors who were exclusively involved in data acquisition and critical revision of the manuscript are listed following alphabetical order in the authorship list.

## Additional Contributions

We would like to thank Hyemin Han, Educational Psychology Program, University of Alabama, Tuscaloosa, AL, USA, for his help with the BayesFactorFMRI toolbox.

