## Supplemental Information for "Are Brain Responses to Emotion a Reliable Endophenotype of Schizophrenia? An Image-based fMRI Meta-analysis"

Fiorito *et al.*

**Contents:**

Supplementary Methods

Supplementary Tables S1-S3

Supplementary Figure S1

Supplementary References

**Supplementary Methods**

*Study exclusion*

27 studies were excluded due to the following reasons:

- 8 studies employed the same dataset as in another included study(1–8)
- 8 studies used non-relevant task design (1 non-visual stimuli(9), 1 non-visual stimuli and conditioning task(10), 1 positively valenced emotional stimuli(11), 1 Theory of Mind task(12,13), 1 stress induction task(14), 1 rejection-acceptance task(15), 1 Stroop task using words(16))
- 4 studies were conference abstracts(17–20)
- 3 studies did not perform group comparisons(21–23)
- 2 studies were systematic reviews or meta-analyses(24,25)
- 1 study was a methods paper(26)
- 1 study provided dubious data(27). Reasons for questioning the reliability of this study were that within-group T-maps showed highly unexpected results in healthy controls, opposite to what is typically observed in studies of emotional processing; unfortunately, the authors declined to engage in double-checking their results when we invited them to, so we preferred to exclude this study

*Criteria for contrast selection*

When more than one article was published using the same sample of participants, we favored the article using methods most closely aligned with our inclusion criteria (e.g., we discarded studies that only report region of interest (ROI) results in favor of studies that report whole-brain analyses), and then we favored the one that could provide the contrast maximizing homogeneity between studies. When more than one contrast from the same study was available, we chose the contrast expected to maximize amygdala activation (e.g., contrasts related to the implicit rather than explicit condition(28), or, when a contrast with all negative emotions pooled together was not available, a contrast favoring fearful stimuli over other emotions(29)). Finally, in keeping with the goal of specifically isolating emotional processing regardless of content, when possible we preferred contrasts of the form negative emotional stimuli versus neutral stimuli, rather than negative emotional stimuli versus control condition/implicit baseline.

*Partial brain coverage inclusion*

There is a common consensus regarding the need to exclude studies only employing ROI analyses, in order to avoid biasing whole-brain results in favor of these regions(30). In this meta-analysis, ROI studies were thus excluded. However, we included one study that used a thick-slab acquisition -and thus had a partial brain coverage(31)- given that this coverage was much wider than classical ROIs and encompassed the main brain regions typically involved in emotion processing such as the amygdala (see Supplementary Figure S1 in Wolf et al., 2011 for an image of the thick-slab acquisition employed).

Also, it is important to note that, since the latest version of SDM (SDM-PSI 6.21) no longer assumes a uniform distribution of false positive foci and does not test for spatial convergence, inclusion of studies with partial brain coverage does not necessarily lead to increased Type I errors. On the contrary, this could increase Type II errors in regions not covered by these studies(32).

*Heterogeneity, Publication Bias and Robustness*

I² statistic, which represents the percentage of total variation due to between-study heterogeneity rather than sampling error, was visually inspected at the whole-brain level through the I^2^ statistic heterogeneity map (a meta-analytical map storing for each voxel a value of I^2^ statistic). Heterogeneity is considered to be low when I² values are below 40%, moderate when I² values are between 30-60%, substantial when I² values are between 50-90%, and considerable when I² values are between 75-100%(33). The inspection of the heterogeneity map suggests the presence of low or moderate heterogeneity almost across the whole brain, with only few clusters indicating substantial heterogeneity (see Figure S1). Moreover, I^2^ statistic values were extracted from amygdala ROIs with SDM-PSI. The presence of low heterogeneity in the amygdala was suggested by low I² statistics in the left (I²=10.6%) and the right (I²=15.5%) amygdala. This was also reflected by all included studies lying inside the triangular region of the funnel plot (see Figure 4C in the main text).

Additional analyses were conducted to explore the possible importance of between-study heterogeneity on the results. In order to determine whether results differ according to the definition of “at risk of schizophrenia” (i.e., familial risk, clinical high risk, or psychometric risk), we performed three whole-brain subgroup meta-analyses. The effect of the type of contrast (i.e., negative emotional versus neutral stimuli contrast, or negative emotional versus control condition/implicit baseline contrast) as well as the effect of the task itself (i.e., implicit or explicit task) were also addressed through whole-brain subgroup meta-analyses. We also ran an additional analysis only including studies employing faces, in order to further homogenize the stimuli used across several tasks. Finally, a whole-brain meta-regression with a linear model using participants’ mean age as a regressor was performed to see whether age differences across studies influenced the results. Due to the limited number of studies included in subgroup analyses, and in order to reduce the increased risk of Type I error associated with multiple tests, results of additional analyses were thresholded at p_TFCE_<0.005 as previously done in Dugré et al., 2020(34). No statistically significant results were found in either of the above sensitivity analyses.

Publication bias was assessed in amygdala ROIs, first through visual inspection of the funnel plot, which represents precision of each study as a function of its effect size. In the absence of publication bias, studies are expected to be symmetrically distributed (see Figure 4C in the main text for visual inspection of publication bias). Secondly, we used Egger’s regression test, a quantitative method that tests for the presence of asymmetry in the funnel plot, which was not significant for either left (z=-0.02, p=0.98) or right (z=-0.65, p=0.52) amygdala, indicating reasonable symmetry of the funnel plot and thus no evidence of a publication bias.

Finally, we performed sensitivity analyses for the purpose of examining the robustness of results and identifying outlier studies. For this analysis, we used a jackknife procedure, consisting of discarding one whole-brain T-map of the meta-analysis dataset at a time. The lack of significant differences in brain activation in at-risk individuals compared with healthy controls was replicated in all whole-brain jackknife analyses, suggesting that this lack of significance was not driven by single outlier studies.

*Partial coverage of the amygdala*

The proximity of the amygdala to the sphenoid sinus makes the BOLD signal in this region more vulnerable to susceptibility artifacts caused by air-tissue interface(35). The resulting dropout of the BOLD signal in this region led to partial coverage of the amygdala in several studies included in this meta-analysis(36–40). In order to limit the influence of this partial coverage at the group-level, the authors of these studies were contacted and asked to rerun their analyses after modifying an SPM default parameter (all studies used SPM). Indeed, in order to create first-level brain masks, SPM employs a default threshold that restricts the statistical analyses only to voxels that exhibit a value that is at least 80% of the mean global signal present in the data. If there is signal dropout in at least one participant, for instance due to susceptibility artifacts, the second-level mask (which corresponds to the intersection of first-level masks) will not cover this region. Therefore, authors were asked to rerun analysis after changing the default threshold parameter from 80% to 20%, in order to enlarge first-level brain masks, while also applying an explicit mask excluding voxels outside of the brain.

Two authors successfully engaged in this process and could provide T-maps with an improved coverage of the amygdala(36,40). Three studies with a partial amygdala coverage were still included since authors could not perform the requested analysis due to time restrictions(37–39).

Future studies, and particularly those targeting commonly artefacted regions like the amygdala, should employ acquisition protocols that counteract the negative consequences of these artifacts(41).

**Supplementary Tables**

**Table S1. PRISMA Checklist**

| **Section and Topic** | **Item #** | **Checklist item** | **Location where item is reported**  **Page #** |
| --- | --- | --- | --- |
| **TITLE** | | |  |
| Title | 1 | Identify the report as a systematic review. | 1 |
| **ABSTRACT** | | |  |
| Abstract | 2 | See the PRISMA 2020 for Abstracts checklist. | 4 |
| **INTRODUCTION** | | |  |
| Rationale | 3 | Describe the rationale for the review in the context of existing knowledge. | 5-6 |
| Objectives | 4 | Provide an explicit statement of the objective(s) or question(s) the review addresses. | 7 |
| **METHODS** | | |  |
| Eligibility criteria | 5 | Specify the inclusion and exclusion criteria for the review and how studies were grouped for the syntheses. | 8-9 |
| Information sources | 6 | Specify all databases, registers, websites, organisations, reference lists and other sources searched or consulted to identify studies. Specify the date when each source was last searched or consulted. | 8 |
| Search strategy | 7 | Present the full search strategies for all databases, registers and websites, including any filters and limits used. | 8 |
| Selection process | 8 | Specify the methods used to decide whether a study met the inclusion criteria of the review, including how many reviewers screened each record and each report retrieved, whether they worked independently, and if applicable, details of automation tools used in the process. | 9, Supp Methods |
| Data collection process | 9 | Specify the methods used to collect data from reports, including how many reviewers collected data from each report, whether they worked independently, any processes for obtaining or confirming data from study investigators, and if applicable, details of automation tools used in the process. | 9 |
| Data items | 10a | List and define all outcomes for which data were sought. Specify whether all results that were compatible with each outcome domain in each study were sought (e.g. for all measures, time points, analyses), and if not, the methods used to decide which results to collect. | 8, Supp Methods |
|  | 10b | List and define all other variables for which data were sought (e.g. participant and intervention characteristics, funding sources). Describe any assumptions made about any missing or unclear information. | 10, Table 1, Table S2 |
| Study risk of bias assessment | 11 | Specify the methods used to assess risk of bias in the included studies, including details of the tool(s) used, how many reviewers assessed each study and whether they worked independently, and if applicable, details of automation tools used in the process. | NA |
| Effect measures | 12 | Specify for each outcome the effect measure(s) (e.g. risk ratio, mean difference) used in the synthesis or presentation of results. | Figure 4 |
| Synthesis methods | 13a | Describe the processes used to decide which studies were eligible for each synthesis (e.g. tabulating the study intervention characteristics and comparing against the planned groups for each synthesis (item #5)). | 8-9 |
|  | 13b | Describe any methods required to prepare the data for presentation or synthesis, such as handling of missing summary statistics, or data conversions. | NA |
|  | 13c | Describe any methods used to tabulate or visually display results of individual studies and syntheses. | 14-15 |
|  | 13d | Describe any methods used to synthesize results and provide a rationale for the choice(s). If meta-analysis was performed, describe the model(s), method(s) to identify the presence and extent of statistical heterogeneity, and software package(s) used. | 12-15, Supp Methods |
|  | 13e | Describe any methods used to explore possible causes of heterogeneity among study results (e.g. subgroup analysis, meta-regression). | Supp Methods |
|  | 13f | Describe any sensitivity analyses conducted to assess robustness of the synthesized results. | Supp Methods |
| Reporting bias assessment | 14 | Describe any methods used to assess risk of bias due to missing results in a synthesis (arising from reporting biases). | Supp Methods |
| Certainty assessment | 15 | Describe any methods used to assess certainty (or confidence) in the body of evidence for an outcome. | Supp Methods |
| **RESULTS** | | |  |
| Study selection | 16a | Describe the results of the search and selection process, from the number of records identified in the search to the number of studies included in the review, ideally using a flow diagram. | 9, Figure 1 |
|  | 16b | Cite studies that might appear to meet the inclusion criteria, but which were excluded, and explain why they were excluded. | Supp Methods |
| Study characteristics | 17 | Cite each included study and present its characteristics. | Table 1 |
| Risk of bias in studies | 18 | Present assessments of risk of bias for each included study. | NA |
| Results of individual studies | 19 | For all outcomes, present, for each study: (a) summary statistics for each group (where appropriate) and (b) an effect estimate and its precision (e.g. confidence/credible interval), ideally using structured tables or plots. | Figure 4 |
| Results of syntheses | 20a | For each synthesis, briefly summarise the characteristics and risk of bias among contributing studies. | Table 1, Table S2, Figure 4 |
|  | 20b | Present results of all statistical syntheses conducted. If meta-analysis was done, present for each the summary estimate and its precision (e.g. confidence/credible interval) and measures of statistical heterogeneity. If comparing groups, describe the direction of the effect. | 16-20, Figure S1 |
|  | 20c | Present results of all investigations of possible causes of heterogeneity among study results. | 18, Supp Methods |
|  | 20d | Present results of all sensitivity analyses conducted to assess the robustness of the synthesized results. | 19, Supp Methods |
| Reporting biases | 21 | Present assessments of risk of bias due to missing results (arising from reporting biases) for each synthesis assessed. | Figure 4, Supp Methods |
| Certainty of evidence | 22 | Present assessments of certainty (or confidence) in the body of evidence for each outcome assessed. | 18 |
| **DISCUSSION** | | |  |
| Discussion | 23a | Provide a general interpretation of the results in the context of other evidence. | 21-22 |
|  | 23b | Discuss any limitations of the evidence included in the review. | 22-23, Supp Methods |
|  | 23c | Discuss any limitations of the review processes used. | 22-23 |
|  | 23d | Discuss implications of the results for practice, policy, and future research. | 24-25 |
| **OTHER INFORMATION** | | |  |
| Registration and protocol | 24a | Provide registration information for the review, including register name and registration number, or state that the review was not registered. | 8 |
|  | 24b | Indicate where the review protocol can be accessed, or state that a protocol was not prepared. | 8 |
|  | 24c | Describe and explain any amendments to information provided at registration or in the protocol. | NA |
| Support | 25 | Describe sources of financial or non-financial support for the review, and the role of the funders or sponsors in the review. | 26-27 |
| Competing interests | 26 | Declare any competing interests of review authors. | 27 |
| Availability of data, code and other materials | 27 | Report which of the following are publicly available and where they can be found: template data collection forms; data extracted from included studies; data used for all analyses; analytic code; any other materials used in the review. | 16 |

*From:* Page MJ, McKenzie JE, Bossuyt PM, Boutron I, Hoffmann TC, Mulrow CD, et al. The PRISMA 2020 statement: an updated guideline for reporting systematic reviews. BMJ 2021;372:n71. doi: 10.1136/bmj.n71. For more information, visit: <http://www.prisma-statement.org/>

**Table S2. Demographic and methodological characteristics of included studies**

| **Source** | **Age, Mean (SD)** | **Male Sex, No. (%)** | **MRI Field strength** |
| --- | --- | --- | --- |
| Bourque et al., 2017(42) | At risk: 14.26 (0.3);  HC: 14.35 (0.4) | At risk: 10 (35.7);  HC: 47 (34.8) | 3T |
| Cao et al., 2016(43) | At risk: 33.29 (12.6);  HC: 32.69 (10.1) | At risk: 20 (34.5);  HC: 39 (41.5) | 3T |
| de Achával et al., 2012(40) | At risk: 30.4 (4.8);  HC: 28.4 (8.3) | At risk: 8 (57.1);  HC: 8 (57.1) | 3T |
| Diwadkar et al., 2012(36) | At risk: 14.3 (3.1);  HC: 14.6 (2.6) | At risk: 12 (63.2);  HC: 16 (66.7) | 4T |
| Modinos et al., 2010(44) | At risk: 19.8 (1.8);  HC: 21 (2.8) | At risk: 7 (41.2);  HC: 7 (41.2) | 3T |
| Modinos et al., 2015(37) | At risk: 24.4 (4.1);  HC: 23.8 (4.6) | At risk: 10 (55.6);  HC: 10 (45) | 1.5T |
| Modinos et al., 2017(39) | At risk: 27.36 (7.6);  HC: 27 (5.6) | At risk: 10 (47.6);  HC: 13 (59.1) | 3T |
| Park et al., 2016(45) | At risk: 23.9 (5.6);  HC: 23.06 (3.9) | At risk: 7 (35);  HC: 8 (47.1) | 3T |
| Pulkkinen et al., 2015(46) | At risk: 22.4 (0.8);  HC: 22.3 (0.7) | At risk: 20 (39.2);  HC: 20 (38.5) | 1.5T |
| Quarto et al., 2018(47) | At risk: 35.4 (10.1);  HC: 31.4 (10.4) | At risk: 13 (36.1);  HC: 30 (53.6) | 3T |
| Rasetti et al., 2009(48) | At risk: 34.8(1.8);  HC: 31.8 (2.2) | At risk: 16 (55.2);  HC: 15 (75) | 3T |
| Spilka et al., 2015(49) | At risk: 41.19 (15.5);  HC: 40.7 (11.1) | At risk: 10 (37);  HC: 13 (48) | 3T |
| van der Velde et al., 2015(38) | At risk: 23.1 (4.4);  HC: 22.1 (3.6) | At risk: 8 (53);  HC: 8 (50) | 3T |
| Shivakumar et al., in preparation(50) | At risk: 29 (1.7);  HC: 31 (4.8) | At risk: 12 (92.3);  HC: 11 (73.3) | 3T |
| Wang et al., 2018(51) | At risk: 19.21 (0.9);  HC: 19.23 (0.9) | At risk: 17 (50);  HC: 13 (43.3) | 3T |
| Wolf et al., 2011(31) | At risk: 42.3 (14.8);  HC: 39 (10.7) | At risk: 9 (45);  HC: 12 (48) | 3T |
| Wolf et al., 2015(52) | At risk: 15.7 (2.7);  HC: 16.6 (3.0) | At risk: 123 (47.3);  HC: 104 (47.3) | 3T |

Abbreviations: HC, Healthy Controls

**Table S3. MNI coordinates of within-group frequentist meta-analysis**

We employed the Hammersmith brain atlas (n30r83, © Copyright Imperial College of Science, Technology and Medicine 2007. All rights reserved(53)) in order to determine the name of brain structures

| **Brain region** | **Hemisphere** | **MNI (x,y,z)** | **SDM-Z** |
| --- | --- | --- | --- |
| ***Healthy controls - activations*** | | | |
| Posterior/lateral orbital gyrus | Right | 36,28,-8 | 4.323 |
| Anterior medial temporal lobe | Right | 38,6,-32 | 4.310 |
| Inferior frontal gyrus | Right | 48,30,16 | 4.076 |
| Thalamus | Right | 6,-6,4 | 4.043 |
| Thalamus | Left | -4,-10,4 | 4.000 |
| Posterior/lateral orbital gyrus | Left | -40,42,-10 | 4.394 |
| Middle frontal gyrus | Left | -36,34,14 | 4.010 |
| Inferior frontal gyrus | Left | -42,28,-8 | 3.879 |
| Amygdala | Left | -22,-6,-18 | 3.698 |
| Anterior part of superior temporal gyrus | Left | -44,16,-26 | 3.625 |
| Cerebellum | Right | 32,-80,-32 | 4.875 |
| Posterior temporal lobe | Right | 38,-62,-20 | 4.420 |
| Lateral occipital lobe | Right | 40,-72,-14 | 3.932 |
| Cerebellum | Left | -36,-48,-34 | 4.431 |
| Lateral occipital lobe | Left | -32,-80,-14 | 4.325 |
| Temporal horn of lateral ventricle | Left | -30,-22,-10 | 2.800 |
| Amygdala | Right | 22,-4,-18 | 3.299 |
| ***Healthy controls - deactivations*** | | | |
| Postcentral gyrus | Right | 50,-14,16 | -5.161 |
| Insula | Right | 44,-6,4 | -5.057 |
| Posterior superior temporal gyrus | Right | 64,-12,6 | -5.043 |
| Inferiolateral parietal lobe | Right | 50,-24,22 | -3.832 |
| Posterior superior temporal gyrus | Left | -62,-24,8 | -5.859 |
| Postcentral gyrus | Left | -54,-16,14 | -5.647 |
| Inferiolateral parietal lobe | Left | -44,-26,22 | -4.821 |
| Insula | Left | -38,-20,6 | -3.826 |
| Precentral gyrus | Right | 8,-24,52 | -4.053 |
| ***At risk - activations*** | | | |
| Cerebellum | Left | -28,-78,-32 | 5.776 |
| Lateral occipital lobe | Left | -42,-68,-16 | 5.404 |
| Lateral occipital lobe | Right | 40,-66,-18 | 5.244 |
| Cerebellum | Right | 38,-56,-26 | 4.398 |
| Thalamus | Right | 4,-16,12 | 4.828 |
| Thalamus | Left | -14,-16,18 | 4.599 |
| Caudate nucleus | Right | 12,6,8 | 4.496 |
| Insula | Right | 32,-6,-18 | 4.309 |
| Posterior orbital gyrus | Left | -26,10,-16 | 4.147 |
| Inferior frontal gyrus | Left | -48,36,-8 | 5.107 |
| Middle frontal gyrus | Left | -48,46,8 | 5.021 |
| Inferior frontal gyrus | Right | 44,20,16 | 6.422 |
| Anterior part of superior temporal gyrus | Right | 52,18,-16 | 4.906 |
| Middle frontal gyrus | Right | 32,8,30 | 4.431 |
| Superior frontal gyrus | Right | 2,16,54 | 4.848 |
| Superior frontal gyrus | Left | -6,32,50 | 4.704 |
| Posterior temporal lobe | Right | 52,-42,-2 | 3.713 |
| Amygdala | Left | -28,0-19 | 3.763 |
| Amygdala | Right | 26,0,-19 | 3.713 |
| ***At risk - deactivations*** | | | |
| Postcentral gyrus | Right | 62,-6,8 | -5.317 |
| Posterior superior temporal gyrus | Right | 62,-6,4 | -5.105 |
| Middle inferior temporal gyrus | Right | 62,-16,-22 | -4.389 |
| Pre-subgenual frontal cortex | Left | -10,32,-10 | -5.094 |
| Superior frontal gyrus | Right | 4,46,-4 | -4.482 |

p_TFCE_<0.05

**Supplementary Figure**

**Figure S1. Whole-brain heterogeneity map**

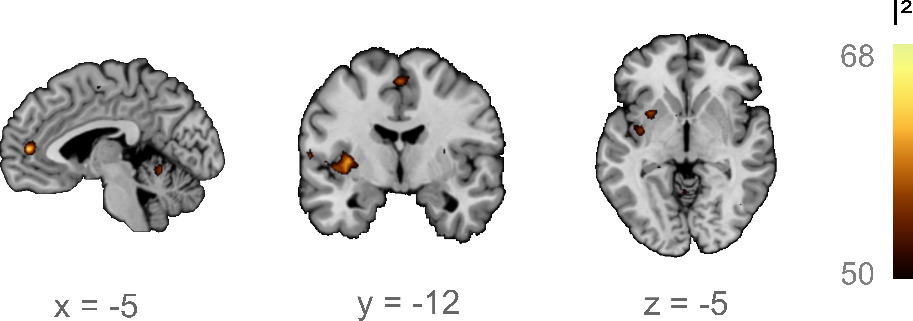

*Heterogeneity map displaying for each voxel the I² statistic. Since an I² statistic above 50% is commonly interpreted as substantial heterogeneity, the map was thresholded at I²>50%. A few small clusters displayed evidence of substantial heterogeneity in bilateral superior frontal gyrus, cerebellum, posterior temporal lobe, substantia nigra and left insula. Functional T-maps are overlaid on the Colin 27 anatomical template.*
